# SARS-CoV-2 seroprevalence and neutralizing activity in donor and patient blood from the San Francisco Bay Area

**DOI:** 10.1101/2020.05.19.20107482

**Authors:** Dianna L. Ng, Gregory M. Goldgof, Brian R. Shy, Andrew G. Levine, Joanna Balcerek, Sagar P. Bapat, John Prostko, Mary Rodgers, Kelly Coller, Sandy Pearce, Sergej Franz, Li Du, Mars Stone, Satish K. Pillai, Alicia Sotomayor-Gonzalez, Venice Servellita, Claudia Sanchez San Martin, Andrea Granados, Dustin R. Glasner, Lucy M. Han, Kent Truong, Naomi Akagi, David N. Nguyen, Neil M. Neumann, Daniel Qazi, Elaine Hsu, Wei Gu, Yale A. Santos, Brian Custer, Valerie Green, Phillip Williamson, Nancy K. Hills, Chuanyi M. Lu, Jeffrey D. Whitman, Susan Stramer, Candace Wang, Kevin Reyes, Jill M.C. Hakim, Kirk Sujishi, Fariba Alazzeh, Lori Pham, Ching-Ying Oon, Steve Miller, Theodore Kurtz, John Hackett, Graham Simmons, Michael P. Busch, Charles Y. Chiu

**Affiliations:** Department of Laboratory Medicine, University of California, San Francisco, San Francisco, CA, USA; Department of Pathology, University of California, San Francisco, San Francisco, CA, USA; Applied Research and Technology, Abbott Diagnostics, Abbott Park, IL, USA; UCSF-Abbott Viral Diagnostics and Discovery Center, San Francisco, CA, USA; Vitalant Research Institute, San Francisco, CA, USA; Department of Medicine, Division of Infectious Diseases, University of California, San Francisco, San Francisco, CA, USA; Creative Testing Solutions, Tempe, AZ, USA; Department of Neurology, University of California, San Francisco, San Francisco, CA, USA; Department of Epidemiology and Biostatistics, University of California, San Francisco, San Francisco, CA, USA; Lab Medicine Service, San Francisco VA Healthcare System; American Red Cross, Gaithersburg, MD, USA; Department of Medicine at ZSFG, the Division of HIV, ID & Global Medicine

## Abstract

We report very low SARS-CoV-2 seroprevalence in two San Francisco Bay Area populations. Seropositivity was 0.26% in 387 hospitalized patients admitted for non-respiratory indications and 0.1% in 1,000 blood donors. We additionally describe the longitudinal dynamics of immunoglobulin-G, immunoglobulin-M, and *in vitro* neutralizing antibody titers in COVID-19 patients. Neutralizing antibodies rise in tandem with immunoglobulin levels following symptom onset, exhibiting median time to seroconversion within one day of each other, and there is >93% positive percent agreement between detection of immunoglobulin-G and neutralizing titers.

---

Coronavirus disease 2019 (COVID-19) is a novel respiratory illness caused by the severe acute respiratory syndrome coronavirus 2 (SARS-CoV-2)^1^. The symptoms of COVID-19 range from asymptomatic infection to acute respiratory distress syndrome and death, and the COVID-19 pandemic has resulted in substantial burdens on healthcare systems worldwide^2,3^. Given the current state of diagnostic testing which largely relies on molecular techniques, the seroprevalence of SARS-CoV-2-specific antibodies in different populations remains unclear. Accurate and large-scale serologic testing that includes detection of neutralizing antibodies is essential in evaluating spread of infection in the community, informing public health containment efforts, and identifying donors for convalescent plasma therapy trials.

## Performance Characteristics of the Abbott Architect IgG and IgM SARS-CoV-2 Assays

We first assessed the performance of the Abbott Architect SARS-CoV-2 IgG (FDA Emergency Use Authorization (EUA)) and IgM (prototype) assays from a cohort of five outpatients and 38 hospitalized patients at University of California, San Francisco (UCSF) Medical Center and the San Francisco Veterans Affairs (SFVA) Health Care System. These assays are chemiluminescent microparticle immunoassays that target the nucleocapsid and spike proteins, respectively. All patients received care at adult inpatient units or clinics and were RT-PCR positive for SARS-CoV-2 from nasopharyngeal and/or oropharyngeal swab testing (**Figure 1A, Table S1**). The percentage of patients seroconverting for IgG at weekly time intervals following reported symptom onset reached 94.4% at ≥22 days (**Figure 1B**). Correspondingly, IgG assay sensitivity from analysis of all 423 samples increased weekly to reach 96.9% at ≥22 days, and was 99% when samples from seven immunocompromised patients (see below) were excluded (**Figure 1D, Table 1**). The percentage of patients seroconverting for IgM was also 94.4% at ≥22 days (**Figure 1E**) and IgM assay sensitivity from analysis of 346 samples was 97.9% (98.9% with immunocompromised patients excluded) (**Figure 1G, Table 1**).

**Figure 1:**
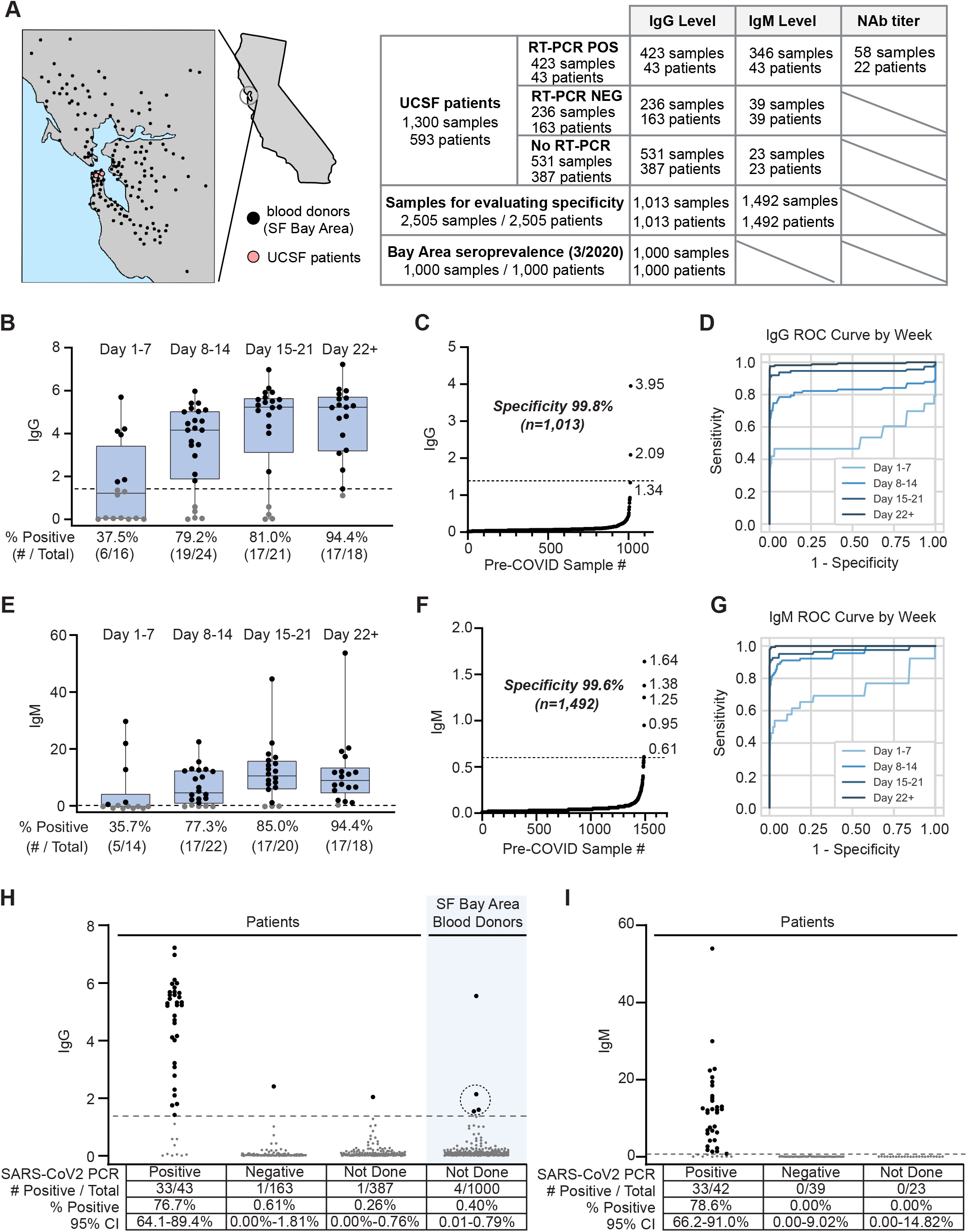
Seroprevalence of Antibodies to SARS-CoV-2. **(A)** Schematic of testing performed and location of patient populations assessed. **(B)** IgG S/C ratios for SARS-CoV-2 PCR-positive patient samples for the indicated weekly timeframes post-onset of symptoms (if multiple samples per patient were collected, the sample with the highest S/C value within each time frame is plotted). The percent of patients with positive antibody responses measured within each timeframe is indicated below the graphs. **(C)** IgG S/C ratios measured in pre-COVID samples; specificity and number of samples is indicated on graph. **(D)** Receiver operating characteristic (ROC) curves for IgG levels for all samples from SARS-CoV-2 PCR-positive patients within the indicated weekly time frames. AUCs for are 0.537 (day 1-7), 0.827 (day 8-14), 0.946 (day 15-21), 0.990 (day 22+). **(E)** IgM S/C ratios, as in **(B)**. **(F)** IgM S/C ratios measured in pre-COVID samples. **(G)** ROC curves for IgM levels, as in **(D)**; AUCs are 0.720 (day 0-7), 0.955 (day 8-14), 0.970 (day 15-21), 0.999 (day 22+). IgG **(H)** and IgM **(I)** S/C ratios were determined for hospitalized patients and outpatients and blood donors on whom SARS-CoV-2 PCR testing was positive or negative or was not performed. Numbers of seroreactive and total individuals tested are shown in tables below the graphs. The circled data points in **(H)** were additionally tested by the VITROS and neutralization assays. For patients with multiple samples, the single highest S/C value is plotted. In (**B**), (**C**), and (**H**), the dotted line at 1.4 indicates cutoff for IgG positivity; in (**E**), (**F**), and (**I**), the dotted line at 0.6 indicates cutoff for IgM positivity; data points in black and gray are above and below the indicated cutoffs, respectively.

**Table 1:**
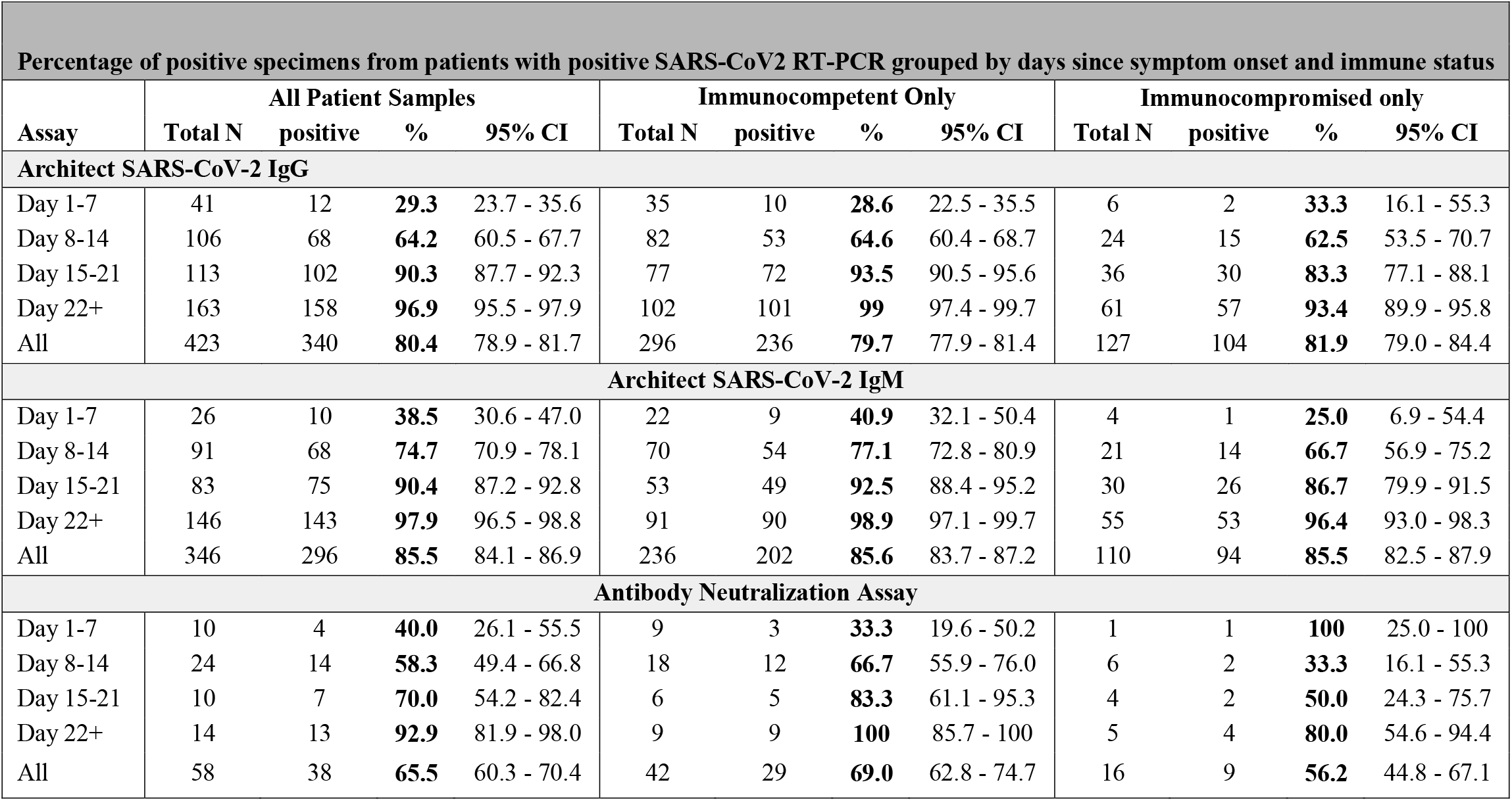
Clinical sensitivities of the Abbott Architect SARS-CoV-2 IgG and IgM and *in vitro* neutralization assays. Clinical sensitivity of each assay, defined as the percent of samples from RT-PCR confirmed SARS-CoV-2 infected patients that test positive in each assay. Total numbers of samples, positive samples, and percent positive among total samples with 95% confidence intervals (CI) are shown for the indicated time frames for samples from all patients (left column), samples from immunocompetent patients only (middle column), and samples from immunocompromised patients only (right column.) Immunocompromised patients: six solid organ transplant recipients on tacrolimus and MMF and one rheumatoid arthritis patient on methotrexate and infliximab.

Of the four patients who had not seroconverted for IgG by the end of 14 days (**Figure 1B**), two were kidney transplant recipients on tacrolimus and mycophenolate mofetil (MMF) immunosuppressive therapy; one was >90 years old; and one was an asymptomatic patient receiving acute psychiatric care who provided an unreliable history. Both renal transplant recipients were observed to ultimately seroconvert for IgG and IgM. Notably, delayed seroconversion for IgG and IgM was not universal in immunosuppressed patients: three additional solid organ transplant (SOT) recipients on tacrolimus and MMF, as well as one patient with rheumatoid arthritis on methotrexate and infliximab, all seroconverted within two weeks. A further SOT recipient was positive for IgG and IgM in the earliest available serum sample from day 17 post symptom onset (**Figure 2D, E**). We did not have samples beyond day 18 for the remaining two patients. However, as seroconversion was observed as late as three weeks after symptom onset (**Figure 2D, E**), it is possible that analysis of later samples would have demonstrated detectable antibodies in their serum. The one patient who was still IgG negative in the 22+ day time frame (**Figure 1B**) (from a plasma sample collected on day 29) had only mild symptoms and was positive by IgM and neutralizing antibody testing (described below). Conversely, the one patient who was IgM negative in the 22+ day time frame was both IgG and neutralizing antibody positive from a plasma sample collected on day 50 (**Figure 1E**), by whichh time IgM antibody titers may have waned significantly.

**Figure 2:**
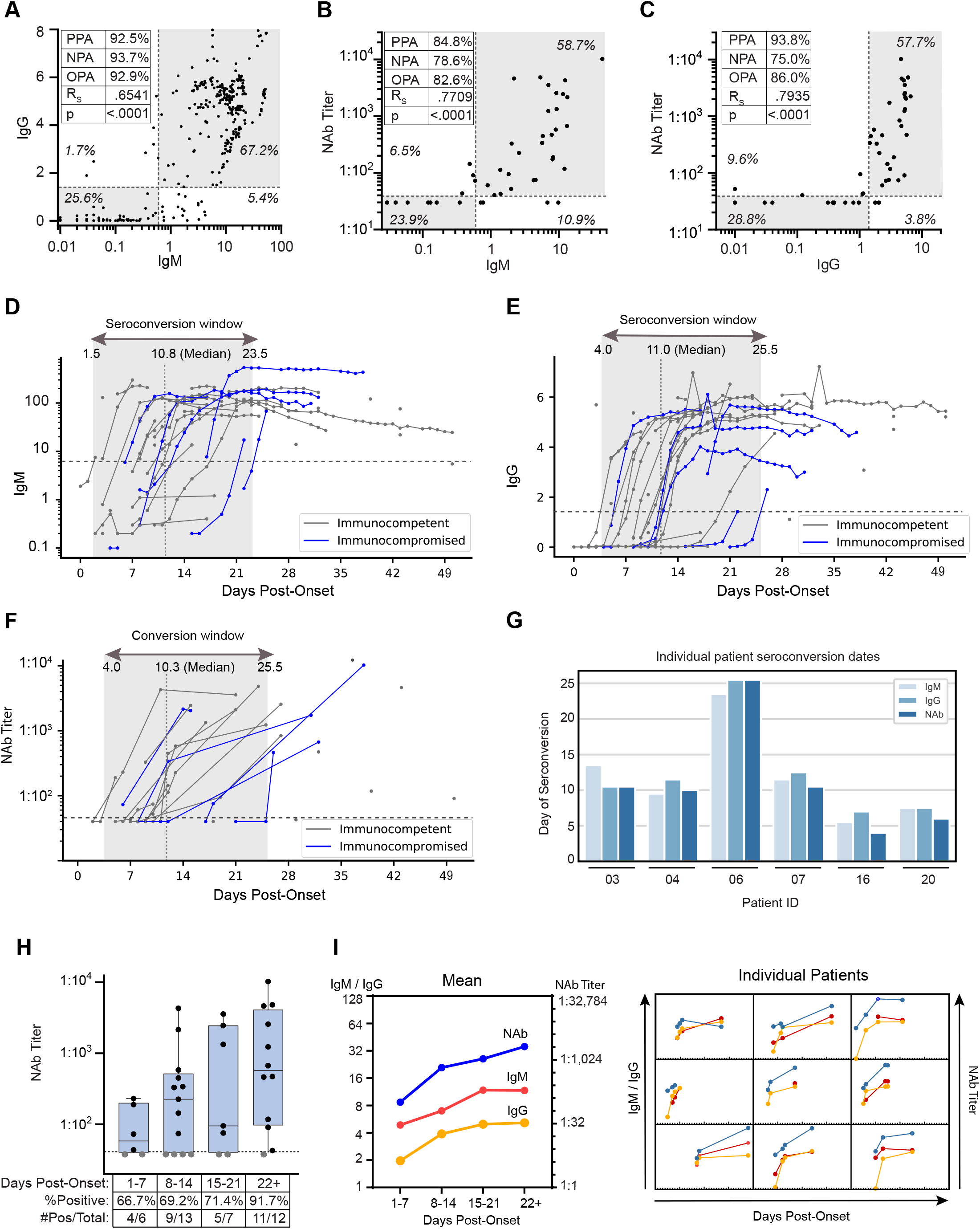
Longitudinal dynamics and *in vitro* neutralizing activity of antibodies against SARS-CoV-2. (**A**) IgG and IgM levels for SARS-CoV-2 PCR positive matched patient samples. Percent of data points in each quadrant and positive percent agreement (PPA), negative percent agreement (NPA), and overall percent agreement (OPA) between IgG and IgM are shown. 80% neutralization titers (NT80) plotted against IgM (**B**) and IgG (**C**) S/C values. The cutoff for NT80 was a titer level of >40; negative results are non-numeric (<40) and are plotted at 35 for visualization purposes. (**D-F**), IgM (**D**) and IgG (**E**) S/C ratios and NT80 titers (**F**) for SARS-CoV-2 PCR-positive patients were plotted against day post symptom onset. Immunocompromised patients are shown in blue. In (**D, E**), for patients with multiple same-day samples, the sample with the highest S/C value is plotted. (**G**) For the 6 SARS-CoV-2 PCR-positive patients whose IgM, IgG, and NT80 seroconversion events were captured during serial sampling, the days post-symptom onset seroconversion events are compared. (**H**) NT80 activity was evaluated per patient for the indicated time frames post onset of symptoms. The percent of patients with detectable NT80 activity measured within each time frame is indicated below the graphs. If multiple samples per patient were collected, the sample with the highest NT80 value within each time frame was used. (**I**) The average NT80 activity (right axis) and IgG and IgM (left axis) levels are plotted by day post-symptom onset (left); corresponding graphs for individual patients are shown in a 3×3 grid (right). If multiple samples per patient were collected, the sample with the highest S/C or NT80 value per time frame was used.

To evaluate assay specificity, serum and plasma samples collected by Abbott Laboratories from US blood donors prior to the COVID-19 pandemic (pre-COVID-19) were tested for IgG (n=1,013) and IgM (n=1,492) seroreactivity. Two samples out of 1,013 were positive by IgG testing, yielding a specificity of 99.8% (95% CI: 99.3-100%) (**Figure 1C**), concordant with the 99.9% specificity reported in a study by the University of Washington^4,5^. Similarly, testing of 235 remnant plasma samples from 163 SARS-CoV-2 PCR-negative UCSF patients collected from late March to early April 2020 resulted in detection of only one positive sample, yielding a specificity of 99.6% (95% CI: 97.7-100%) (**Figure 1H**). The IgG positive sample was from a patient admitted for syncope but who reported a cough of one-month duration, suggesting a potential prior infection with SARS-CoV-2. Six samples out of 1,492 from US blood donors were positive by IgM testing, yielding a specificity of 99.6% (95% CI: 99.2-99.9%) (**Figure 1F**). This was consistent with more limited testing of 39 SARS-CoV-2 PCR negative UCSF patients, none of whom were positive for IgM antibody (**Figure 1I**). Thus, the Architect SARS-CoV-2 IgG and IgM assays demonstrated high sensitivity (96.9%-97.9% at ≥22 days in a primarily hospitalized patient cohort) and specificity (99.6-99.8% in pre-COVID blood donors), with good correlation (*rho* = 0.65) between IgG anti-nucleocapsid protein and IgM anti-spike protein seropositivity (**Figure 2A**).

## Seroprevalence of SARS-CoV-2 in blood donors and patients from the San Francisco Bay Area in March 2020

Next, to investigate SARS-CoV-2 seroprevalence in the San Francisco Bay Area, we collected plasma and serum samples from two cohorts of individuals with low suspicion of infection from COVID-19. One cohort consisted of 1,000 individuals who donated blood in March 2020 at blood bank centers throughout the Bay Area (**Figure 1A, Table S2**). Routine blood donor screening was performed to exclude those with self-reported symptoms of acute illness and abnormal vital signs. We detected four IgG positive samples in this cohort, yielding a seroreactivity rate of 0.40% (**Figure 1H**). This cohort was not tested for IgM antibody. We then analyzed the four samples using two orthogonal tests, the VITROS anti-SARS-CoV-2 total antibody assay (Ortho Clinical Diagnostics EUA) and a SARS-CoV-2 pseudovirus neutralization assay (described below). Three of four samples were negative by both the VITROS and neutralization assays, and thus were designated likely false positives by the Architect IgG assay. Thus, the calculated seroprevalence after confirmatory orthogonal testing for Bay Area blood donors in March 2020 was 0.1% (95% CI: 0.00% - 0.56%). The false positive rate in this population of 0.3% is consistent with with the reported specificity of the Architect SARS-CoV-IgG test of 99.6%^4^.

The other cohort for evaluating seroprevalence represented a cross-section of patients who received care at adult inpatient units or clinics at the UCSF Medical Center for indications other than COVID-19 respiratory disease (non-COVID-19, never tested for SARS-CoV-2 by RT-PCR) from late March to early April 2020. Remnant samples from 532 blood draws taken from these 387 patients were obtained from UCSF clinical laboratories. Of these 532 samples, five were positive for IgG; strikingly, all five of these samples were from the same patient who had respiratory failure and ground-glass opacities on chest imaging but was never tested for SARS-2-CoV by RT-PCR (**Figure 1H**). IgG seroprevalence in this population was thus 0.26% (95% CI: 0-0.76%). Although only 23 of the 532 remnant samples were able to be subsequently tested for IgM antibodies, importantly, none were positive (**Figure 1I**).

## Longitudinal dynamics of immunoglobulin and neutralizing antibody titers in SARS-CoV-2 infected patient

We next analyzed the longitudinal dynamics of plasma IgG (286 samples) and IgM (249 samples) levels in our cohort of 43 patients who were positive for SARS-CoV-2 by PCR. As previously reported, IgG and IgM antibody levels were observed to rise approximately in tandem (**Figure 2D, E**)^6–10^. We correlated median IgG, IgM, and neutralizing antibody (described below) levels at the weekly time intervals with severity of disease, and the differences were not statistically significant.

Lastly, we sought to correlate IgG and IgM seropositivity with SARS-CoV-2 *in vitro* neutralizing activity against a SARS-CoV-2 pseudovirus (a vesicular stomatitis virus (VSV) pseudotype expressing the SARS-CoV-2 spike protein). Plasma titers that achieved 80% neutralization of pseudovirus infectivity (NT80) were measured by luciferase assay (see Methods). We compared NT80 with IgG and IgM measurements in 54 available plasma samples from 22 of the 43 SARS-CoV-2 PCR positive patients (**Figure 2B, C**). The positive percent agreement (PPA) between NT80 and IgG positivity was 93.8% and the negative percent agreement (NPA) was 75.0% (**Figure 2C**). Results from the NT80 and IgM comparison were similar, with a PPA of 84.8% and NPA of 78.6% (**Figure 2B**). Importantly, neutralizing titers appeared concomitantly in plasma with IgG and IgM positivity (**Figure 2D-G**), correlated well with IgG (*rho* = 0.79) and IgM (*rho* = 0.77) levels, and increased over time in parallel with the rise of anti-spike IgM and ant-nucleocapsid IgG antibodies (**Figure 2H-I**).

## Conclusions

In this study, we provide evidence that seropositive results using the Architect SARS-CoV-2 anti-nucleocapsid protein IgG and anti-spike IgM assays are generally predictive of *in vitro* neutralizing capacity. This correlation may have particular relevance for recovered COVID-19 patients and the identification of candidate donors to provide blood for convalescent plasma therapy. However, *in vitro* neutralization activity may not confer protective immunity and the efficacy of convalescent plasma therapy for treatment of COVID-19 disease remains to be determined. Our results also show that the seroprevalence of IgG antibodies against SARS-CoV-2 in blood donors and non-COVID-19 patients seen at a tertiary care hospital in the San Francisco Bay Area from March to April 2020 is very low at 0.10% (95% CI: 0.00% - 0.56%). and 0.26% (0.00% - 0.76%), respectively. These seroprevalence rates in two distinct populations in the San Francisco Bay Area are near the specificity limit of the Architect assay, and are far lower than the specificity limits for many lateral flow immunoassays^11^. Our findings contrast with those from other community-based studies that reported higher rates of seropositivity in California^12,13^, and underscore the importance of using a highly accurate test for surveillance studies in low-prevalence populations. They also indicate a very low likelihood of widespread cryptic circulation of SARS-CoV-2 in the Bay Area prior to March 2020, consistent with the low detection rate by direct viral testing of respiratory samples collected during that early time period^14^.

## METHODS

### Study design and Ethics

The study population consisted of patients with available remnant serum and plasma specimens from the clinical laboratories at University of California, San Francisco (UCSF). Samples from patients who were positive or negative by SARS-CoV-2 real-time polymerase chain reaction (RT-PCR) testing of nasopharyngeal, oropharyngeal, and/or pooled nasopharyngeal-oropharyngeal swabs were collected in March – April 2020. Additional samples were collected from randomly selected cohorts of outpatients and hospitalized patients at UCSF during the same time period seen for indications other than COVID-19 respiratory disease (non-COVID). Serum samples from blood donors in the San Francisco Bay Area were collected by Vitalant Research Institute in March 2020. Clinical data for UCSF patients were extracted from electronic health records and entered in a HIPAA (Health Insurance Portability and Accountability Act)-secure REDCap research database. Collected data included demographics, major comorbidities, patient-reported symptom onset date, clinical symptoms and indicators of COVID-19 severity such as admission to the intensive care unit and requirement for mechanical ventilation. This study was approved by the institutional review board (IRB) at UCSF (UCSF IRB #10-02598) as a no-subject contact study with waiver of consent.

### Serologic testing

The Abbott Architect SARS-CoV-2 IgG assay (FDA Emergency Use Authorization (EUA)) and SARS-CoV-2 IgM (prototype) testing was performed using either serum or plasma samples on the Architect instrument according to the manufacturer instructions^4^. These tests are chemiluminescent microparticle immunoassay reactions that target the nucleocapsid protein (IgG assay) or the spike protein (IgM assay) and measure relative light units that are then used to calculate an index value. At a predefined index value threshold of 0.6 signal-to-cutoff (S/C) ratio for IgM seropositivity and 1.4 S/C for IgG for seropositivity, these assays were found to have specificities of 99.6% - 99.8%.

The VITROS anti-SARS-CoV-2 total antibody assay approved under FDA Emergency Use Authorization was performed using either serum or plasma samples at Vitalant Research Institute according to the manufacturer instructions^15^. The test is a chemiluminescent immunoassay that targets the spike protein and measures relative light units that are then used to calculate an index value. At a predefined index value threshold of 1.0 signal-to-cutoff (S/C) ratio for IgG seropositivity, this assay was found to have a sensitivity of 100% (92.7% - 100%) and specificity of 100% (95% CI = 99.1% - 100.0%).

### Production of pseudoviruses for the SARS-CoV-2 neutralization assay

VSVΔG-luciferase-based viruses, in which the glycoprotein (G) gene has been replaced with luciferase, were produced by transient transfection of viral glycoprotein expression plasmids (pCG SARS-CoV-2 Spike, provided courtesy of Stefan Pölhmann^16^, as well as pCAGGS VSV-G or pCAGGS EboGP as controls) or no glycoprotein controls into HEK293T cells by TranslT-2020. Briefly, cells were seeded into 15-cm culture dishes and allowed to attach for 24 hours before transfection with 30 μg expression plasmid per plate. The transfection medium was changed at approximately 16 hours post-transfection. The expression-enhancing reagent valproic acid (VPA) was added to a final concentration of 3.75 mM, and the cells were incubated for three to four hours. The medium was changed again, and the cells were inoculated with VSVΔG-luc virus at a multiplicity of infection (MOI) of 0.3 for four hours before the medium was changed again. At about 24 hours post-infection, the supernatants were collected and cleared of debris by filtration through a 0.45-μm syringe filter.

### Antibody neutralization

HEK293T cells were transfected with human ACE2 and TMPRSS2 by TransIT-2020. After 24 hours cells were plated into black 96-well tissue culture treated plates. Serum or plasma was diluted to 1:20 followed by four subsequent 1:4 dilutions. Per well, 50 μl of pseudovirus harboring either SARS-CoV-2 S, VSV-G or EboGP (adjusted to result in ~10,000 RLU in target cells) was mixed with 50 μl of the respective serum or plasma dilution to give a final series of longitudinal serum or plasma dilutions starting at 1:40 and incubated for one hour at 37°C. Controls included wells with VSVΔG (no envelope), without added serum/plasma, and with serum predetermined to possess or lack neutralizing activity. Subsequently, the 100 μl mix was added to the target cells (performed in duplicate) and cells were incubated for 24 hours at 37°C. Supernatants were then removed, cells were lysed, and luciferase activity was read as per manufacturer instructions. Results were calculated as a percentage of no serum control. Each plate was qualified by lack of infection with the no envelope control, and performance of positive and negative controls. Non-linear regression curves and 80% neutralization titers (NT80) were calculated in GraphPad Prism.

### Statistical analysis

We calculated positive percent agreement (PPA), negative percent agreement (NPA), and overall percent agreement (OPA) between the neutralizing antibody result and IgG, assuming IgG to be the gold standard. We then calculated PPA, NPA, and OPA between the neutralizing antibody result and IgM, assuming IgM to be the gold standard. We calculated 95% exact binomial (Clopper-Pearson) confidence intervals for each proportion. IgG, IgM and NT80 levels were non-normally distributed and were summarized using medians and interquartile ranges. We compared antibody levels to dichotomously-defined clinical characteristics at various time points using Wilcoxon rank sum tests. The correlations between age and IgG, IgM, and NT levels were calculated using Spearman non-parametric correlation coefficients. Statistical calculations were performed using python libraries scipy.stats, sklearn.metrics.auc and statsmodels.stats as well as Stata v15.1 (College Station, TX).

**Table S1:** Baseline demographic characteristics, presenting symptoms, chronic medical conditions, medications, and radiographic findings of 43 SARS-CoV-2 PCR-positive UCSF outpatients and hospitalized patients.

**Table S2:** Descriptive demographic characteristics of individuals who donated blood at San Francisco Bay Area community blood centers (Vitalant Research Institute).

## Data Availability

Raw data used in this study, including de-identified patient metadata and test results, are available upon request.

## Acknowledgments

We thank the patients and their families at UCSF without whom collecting and providing this aggregate data would not have been possible.

## Contributions

C.Y.C. conceived, designed, and supervised the study. D.L.N., and G.M.G. coordinated the study. D.L.N., G.M.G., B.R.S., A.G.L., S.P.B., J.B., and C.Y.C. analyzed data, designed figures, and wrote and edited the manuscript. A.S.G., V.S., C.S.S.M., A.G., D.R.G., E.H., W.G., Y.A.S., C.W., K.R., J.H., F.A., L.P., C.-Y.O., C.M.L. contributed to the collection of clinical specimens. K.S., T.K., and E.C.T. coordinated clinical sample collection and IgG testing. S.M. provided clinical data and facilitated sample collection. L.M.H., K.T., N.A., D.N.N., N.M.N., and D.Q. performed chart review. M.S., B.C., V.G., P.W., M.B., and J.D.W. coordinated blood donor samples and data. S.Z., L.D., G.S., and S.K.P. performed neutralizing antibody assays. J.M.C.H., J.P., M.R., K.C., and S.P. performed IgG and IgM testing and provided data establishing testing characteristics of SARS-CoV-2 IgG and IgM assays. N.K.H. performed biostatistical analysis and review. All authors read the manuscript and agreed to its contents.

